# Neonatal Brain Network Integration Trajectories Predict Neurodevelopment in Congenital Heart Disease

**DOI:** 10.64898/2026.06.06.26355074

**Authors:** Lauren Harasymiw, Amy Kuang, Duan Xu, Aaron Scheffler, Elizabeth George, Shabnam Peyvandi, Patrick McQuillen

**Author notes:** **Corresponding author**: Lauren Harasymiw, Mission Hall 550 16th Street, San Francisco, CA 94158-2711.

## Abstract

**Impact statement:**

- In infants with critical congenital heart disease, perioperative growth in whole-brain network integration was associated with cognitive, language, and motor outcomes in early childhood.
- White matter injury was associated with slower growth in network integration, whereas cardiac physiology subtype was not associated with network development.
- This study extends prior work by showing that a longitudinal measure of neonatal whole-brain network development, rather than a single imaging timepoint, predicts neurodevelopment across multiple domains.
- These findings identify early network development as a potential biomarker of neurodevelopmental risk and resilience, and support future risk-stratification and intervention studies in this population.

**Background:** Infants with critical congenital heart disease (CHD) are at high risk for abnormal brain development and later neurodevelopmental impairment. We hypothesized that the trajectory of perioperative whole-brain network development would predict neurodevelopmental outcomes in early childhood.

**Methods:** This prospective longitudinal cohort of neonates with critical CHD (n = 97) underwent preoperative and/or postoperative brain MRI with diffusion imaging. Whole-brain network measures were derived from structural connectomes. Neurodevelopment was assessed between 1 and 4 years using the Bayley Scales of Infant and Toddler Development.

**Results:** White matter injury was associated with slower perioperative growth in global efficiency (*p* = 0.013), a measure of network integration, whereas cardiac physiology was not associated with network development. Infants with greater perioperative increases in global efficiency had higher cognitive (*p* = 0.001), language (*p* < 0.001), and motor (*p* = 0.008) scores. For each 1–standard deviation increase in the trajectory of global efficiency, cognitive scores increased by 8.2 points (95% CI, 3.64–12.78), independent of brain injury and socioeconomic factors.

**Conclusion:** In infants with critical CHD, longitudinal whole-brain network development was associated with neurodevelopment across multiple domains. Early network development may represent a candidate biomarker of neurodevelopmental risk and resilience in this population.

## Introduction

Children born with critical congenital heart disease (CHD) are at elevated risk for adverse neurodevelopmental outcomes extending from infancy into adulthood^1^. Early structural brain abnormalities are common, including smaller brain volumes beginning in utero and an increased rate of acquired postnatal brain injuries, even before neonatal operation^2,3^, with direct associations to ND outcomes^4–6^. Risk factors for these early structural brain abnormalities are multi-factorial^7^.

Diffusion MRI (dMRI) processed for structural connectivity can quantify developmental changes in the growth and organization of white matter tracts and in whole-brain neural network organization^8,9^. This systems-level approach to quantifying brain maturation may help to more fully explain risk and resiliency factors after acquired brain injury in the perioperative period. Network development has been reported to be delayed in neonates with CHD^10^ and linked with early functional outcomes^11^.

To our knowledge, no prior studies have examined whether longitudinal neonatal neural network development predicts neurodevelopmental outcomes in children with congenital heart disease (CHD). We therefore investigated whole-brain structural connectome development during the neonatal perioperative period in a prospective cohort of infants with critical CHD. We linked these trajectories to neurodevelopmental outcomes in early childhood. We hypothesized that cardiac physiology and brain injury would influence the rate of network development and that greater perioperative increases in network integration would be associated with better motor and language outcomes between 1 and 4 years of age.

## Methods

### Ethics

This study was approved by the University of California, San Francisco Institutional Review Board. Written informed consent was obtained from parents or legal guardians of all participants. The study followed the Strengthening the Reporting of Observational Studies in Epidemiology (STROBE) reporting guideline.

### Study Design

This is a secondary analysis of a prospective, longitudinal cohort study of perioperative structural connectome development and neurodevelopmental outcomes in neonates with critical CHD treated at a single tertiary care center.

### Study population

Eligible patients were enrolled as fetuses or newborns at the University of California, San Francisco, from 2011 to 2025. Inclusion criteria included CHD requiring a neonatal operation (i.e., critical congenital heart disease). Exclusion criteria included preterm delivery (<36 weeks), a confirmed or suspected genetic or malformation syndrome, or contraindications to MRI (e.g., pacemakers). Enrolled subjects underwent brain MRI before and after cardiac surgery and completed multiple neurodevelopmental follow-up visits between 1 and 4 years. Patients diagnosed as having either d-Transposition of the Great Arteries (d-TGA), defined as ventriculoarterial discordance resulting in parallel systemic and pulmonary circulations, or Single Ventricle Physiology (SVP), defined as requiring staged palliation due to the functional presence of a single ventricle (e.g., including hypoplastic left heart syndrome and related variants), who completed ND follow-up data were included in the current study (Figure 1). The study was restricted to these two lesions to minimize heterogeneity related to cardiovascular physiology.

**Figure 1.**
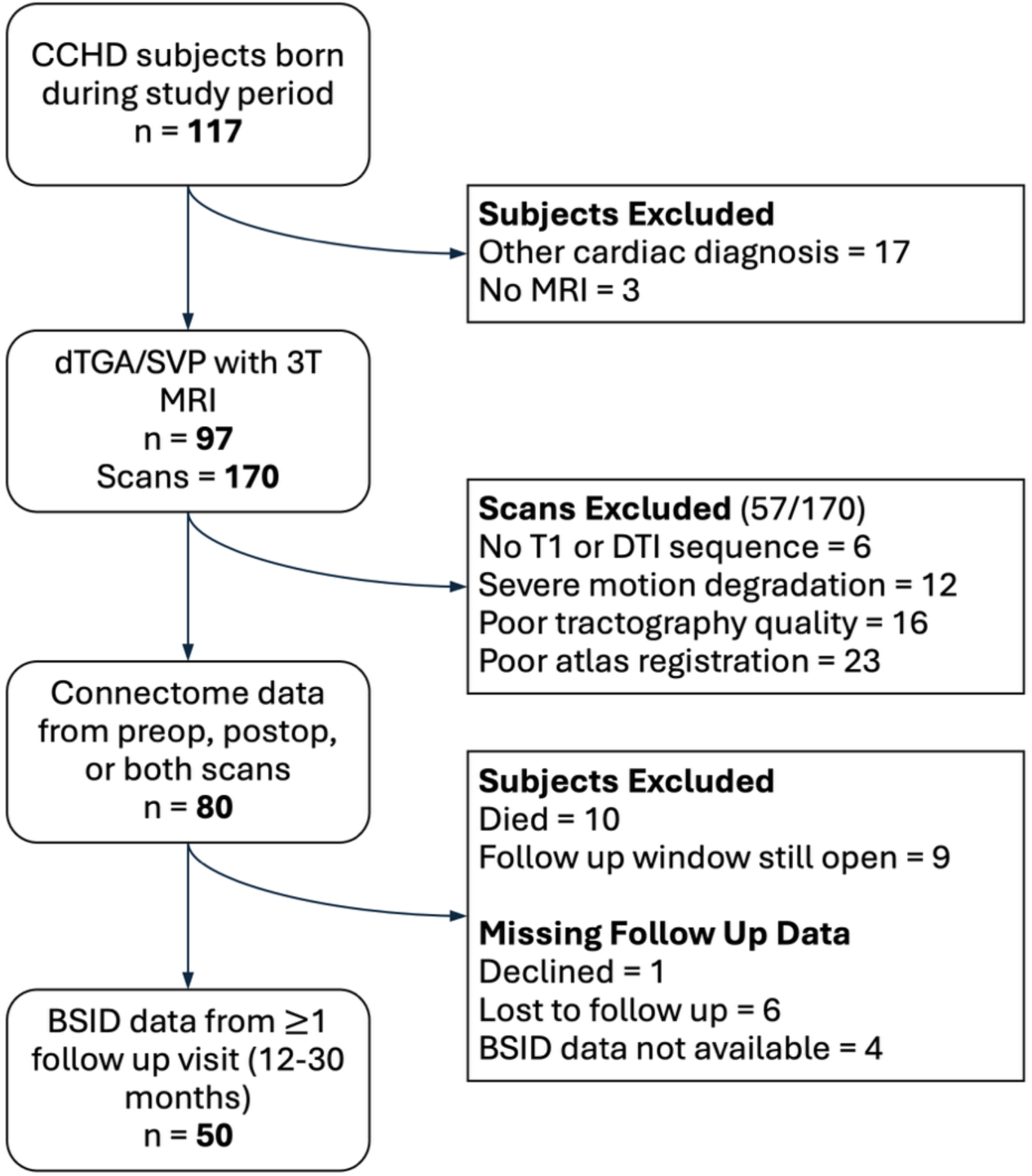
Flow chart of included subjects and data sources. Study period included infants born between November 2011, which was the start of neonatal 3T MRI acquisition at the study site, through January 2025, with the follow-up period extending through August 2025.

### Clinical and sociodemographic data acquisition

Clinical and demographic data, including sex, gestational age at birth and at MRI, birth weight, and race/ethnicity, were abstracted from the electronic medical record. The Neonatal Acute Physiology-Perinatal Extension (SNAPPE) score was used to quantify peri-delivery clinical status. Socioeconomic variables, including maternal education level (years) and neighborhood context (Childhood Opportunity Index state-normalized z-score by zip code), were abstracted from surveys completed by either parent.

### Neurodevelopmental testing

Subjects were invited to complete follow-up neurodevelopmental testing between 1 and 4 years, with visits centered at 12, 18, and 30 months corrected gestational age. Subjects were tested using either the Bayley Scales of Infant & Toddler Development (BSID), Ed. 3 (Bayley-III)^12^ or Ed. 4 (Bayley-4)^13^.

### Brain MRI and structural connectivity processing

Subjects underwent serial brain MRIs, when possible, including preoperative scans in the first week of life before cardiac surgery and postoperative scans before hospital discharge. Of the total sample of eligible infants (n = 97), 85 infants (88%) completed a preoperative MRI, and 85 infants (88%) completed a postoperative MRI. Detailed MRI acquisition parameters and structural connectivity processing have been described previously^11,14^.

In brief, the majority of infants were scanned using a feed-and-swaddle approach, with sedation used when clinically indicated. Ear protection was used for all subjects. MRI scans were performed on a 3-T Discovery MR750 system (GE HealthCare). They included the following sequences: three-dimensional isotropic T1-weighted inversion-recovery spoiled gradient-recalled echo (repetition time msec/echo time msec/inversion time msec, 8.7/3.5/450; section thickness, 1 mm) and diffusion tensor imaging (5000/63; section thickness, 1.8 mm; no gap; in-plane resolution, 2 × 2 mm; 30 directions; b value, 700 sec/mm2). All studies were reviewed by a pediatric neuroradiologist. Acquired brain injuries, including white matter injury (WMI), stroke, intraventricular hemorrhage (IVH), and hypoxic-ischemic injury, were identified using a published scoring system^15^.

We first ran an automated data-rejection algorithm, previously validated in term neonates, to remove motion-corrupted DTI data. Next, we performed whole-brain deterministic tractography using the Diffusion Toolkit. Each brain was parcellated into 90 regions by mapping the UNC Neonatal Infant Atlas to the infant’s T1-weighted SPGR image. Diffusion images were then co-registered to T1 space with FSL’s FMRIB Linear Image Registration Tool (FLIRT), and we visually checked the alignment, excluding any scans that still showed motion corruption. For each infant, we built an undirected connectivity matrix weighted by fractional anisotropy (FA). The network nodes were formed by the atlas ROIs, streamlines connecting ROI pairs formed the edges, and edge weights were the mean FA along each streamline.

Topological metrics were calculated from individual connectomes in MATLAB 2024b (MathWorks) using the functions efficiency_wei, modularity_und, and transitivity_wu in the Brain Connectivity Toolkit. Adjacency matrices for transitivity were normalized. Modularity was analyzed on the log scale to improve distributional fit.

### Statistical analysis

Statistical analyses were performed using R version 4.4.2 (R Foundation for Statistical Computing). All tests were two-sided, and significance was set as *p* < 0.05, unless stated otherwise. Descriptive statistics were summarized as proportions for categorical variables and as mean and standard deviation or median and interquartile range, as appropriate, for continuous variables. Comparisons were conducted using Chi-square tests or Fisher’s exact tests, and either T-tests or Kruskal-Wallis tests for non-parametric continuous variables.

#### Longitudinal whole-brain network development

Network topology metrics (global efficiency, transitivity, modularity) were modeled as a function of postmenstrual age at MRI with a subject-level random intercept for repeated measures using a linear mixed-effects model in the lme4 package of R. Additional covariates known to impact brain development and neurodevelopmental outcomes, including gestational age, biological sex, and maternal education level, were tested in univariate models and included in the final multivariate models when *p* < 0.05. Based on univariate testing, global efficiency models were adjusted for gestational age and maternal education, and modularity models were adjusted for sex. No additional covariates were retained for transitivity models. Separate models were then fit for each network metric and each exposure of interest: cardiac physiology (d-TGA vs SVP), any brain injury (no injury vs any injury), white matter injury, and stroke. The fixed-effects structure for each model included postmenstrual age at MRI, exposure status, the interaction between postmenstrual age and exposure status, and any covariates identified during univariate testing, as specified above. The effect estimate of interest was the exposure-by-time interaction term, which represents the difference in the slope of network development between exposure groups.

#### Linking network development to neurodevelopment

To evaluate whether early brain network development predicted later neurodevelopment, we used a joint modeling approach. This first-stage model used only neonatal MRI data and did not incorporate neurodevelopmental outcome measures. We focused on the trajectory of global efficiency as our predictor of interest because it was the only network metric significantly affected by brain injury. First, we modeled subject-specific trajectories of network efficiency (E) as a function of postmenstrual age at MRI using a Bayesian linear mixed-effects framework to account for subjects with MRI data from only one timepoint. Postmenstrual age at MRI was standardized to z-scores to facilitate interpretation. The model specified a population slope for standardized PMA and subject-level random intercepts and slopes. For infants with a single MRI timepoint, subject-level slopes were partially informed by the population-level trajectory estimated in the mixed-effects model.

We fit the first-stage model in R using the Bayesian Regression Models using Stan (brms) package, using Hamiltonian Monte Carlo (4,000 iterations with 1,000 warm-up iterations, parallelized across 4 cores) and default priors. We used a target acceptance rate of 0.99 and a maximum tree depth of 15. Convergence was assessed using standard Markov chain Monte Carlo diagnostics, including trace plots, R-hat, and effective sample size. Model fit was assessed with posterior predictive checks. We then extracted posterior summaries (means and 95% credible intervals) of each subject’s conditional intercept and slope. These coefficients were transformed back to the raw PMA scale using the empirical mean and standard deviation to obtain interpretable per-subject trajectories. Across the cohort, the median interval between preoperative and postoperative MRI was 20 days; thus, the estimated slope reflects the rate of network integration change during approximately the first three weeks of life.

For the second-stage model examined whether subject-level global efficiency development predicts neurodevelopmental outcomes. For this model, we fit a linear mixed-effects model in R using the nlme package, with a subject-level random intercept and restricted maximum likelihood (REML). Our primary exposure was the individualized trajectory (slope) of global efficiency across the perioperative period, estimated from longitudinal MRI data in the first-stage model. We used each infant’s posterior mean slope estimate, standardized to a z-score, so that effect estimates could be interpreted as a 1-standard-deviation increase in neonatal global efficiency development. Our outcomes of interest were BSID composite scores analyzed as longitudinal outcomes with repeated measurements obtained between 1 and 4 years of age.

Individual multivariate models were fit for each composite score (i.e., cognitive, motor, language). Cumulative brain injury scoring and maternal education level (as an individual-level measure of socioeconomic status, home environment, and parental cognitive abilities) were included *a priori* as known predictors of neurodevelopmental outcomes. Because scores have been reported to vary across Bayley scaled editions^16^, test edition was also included as a covariate in all models. Additional candidate covariates were screened in univariate linear mixed-effects models, with age at BSID assessment as the time variable to model expected developmental change and subject-specific random intercepts to account for within-subject correlation due to repeated measures, as shown in Supplemental Table 2 (Online). Covariates tested included birth weight z-score, gestational age, sex, cardiac diagnosis, and childhood opportunity index z-score (normalized to state), a composite measure of neighborhood-level education, health, and economic factors. Additional metrics of clinical severity, including SNAPPE score and length of stay, were not included in univariate testing due to their substantial overlap with cardiac diagnosis and the potential to reflect downstream aspects of the postnatal course rather than baseline confounding. Variables associated with composite scores at *p* < 0.10 were included in the final multivariable models. Final covariate sets, therefore, differed slightly across the BSID domains, as shown in Table 2.

## Results

### Study Population

A total of 117 neonates were enrolled during the study period (Figure 1). Of the enrolled subjects, 20 neonates did not meet the inclusion criteria for the current study: 3 did not complete an MRI during the perioperative period, and 17 had a cardiac diagnosis other than d-TGA or SVP. Demographic and neonatal clinical data for the remaining cohort of 97 infants are detailed in Table 1. 52.6% had d-TGA (n = 51), and the remaining 47.4% had SVP (n = 46). Within this cohort, 80 subjects had at least one 3T MRI with both T1 and DTI sequences of sufficient quality to create a structural connectome. A total of 62 preoperative and 61 postoperative scans were included in the final analysis, and 43 subjects contributed connectome data from both scans.

**Table 1.** Sample characteristics.

|  | **d-TGA** | **SVP** | **p-value<sup>1</sup>** |
| --- | --- | --- | --- |
| N (%) | 51 (52.6) | 46 (47.4) |  |
| Male, n (%) | 35 (68.6) | 35 (76.1) | 0.41 |
| Race and Ethnicity, n (%) |  |  | 0.37 |
| Non-Hispanic White | 14 (27.5) | 15 (32.6) |  |
| Hispanic or Latino | 23 (45.1) | 21 (45.6) |  |
| Asian | 7 (13.7) | 3 (6.5) |  |
| Other | 6 (11.8) | 3 (6.5) |  |
| Non-Hispanic Black | 1 (2.0) | 4 (8.7) |  |
| Maternal Education, n (%)<sup>2</sup> |  |  | 0.40 |
| Partial high school or less | 6 (12) | 5 (11) |  |
| High school diploma | 12 (24) | 10 (23) |  |
| Partial college or technical training | 5 (10) | 10 (23) |  |
| College graduate | 17 (34) | 9 (20) |  |
| Graduate school | 10 (20) | 10 (23) |  |
| Gestational age, M (SD) | 39.1 (1.0) | 38.6 (0.9) | 0.03 |
| Birth weight (kg), M (SD) | 3.3 (0.5) | 3.2 (0.4) | 0.37 |
| SNAPPE-II score, M (SD)<sup>3</sup> | 12.6 (10.6) | 18.8 (9.2) | 0.01 |
| Septostomy, n (%) | 22 (43.1) | 3 (6.5) | <0.001 |
| Survival to discharge, n (%) | 49 (96.1) | 43 (93.5) | 0.56 |
| Length of stay, days, median (IQR) | 22 (19-30) | 55 (35-78) | <0.001 |

d-Transposition of the Great Arteries (d-TGA), Single Ventricle Physiology (SVP), Score for Neonatal Acute Physiology with Perinatal Extension (SNAPPE-II), day of life (DOL), interquartile range (IQR).
<sup>1</sup>From chi-square, Kruskal-Wallis, or T-tests. <sup>2</sup>Data missing for 3 subjects. <sup>3</sup>Data missing for 30 subjects.

From the subsample of 80 neonates who contributed imaging data, 61 were eligible for follow-up during the study period, and 50 contributed BSID data from at least one follow-up visit. A detailed breakdown of the reasons for MRI scan exclusion and incomplete follow-up is provided in Figure 1.

### Brain Injury

A high rate of brain injuries, including WMI, intraventricular hemorrhage (IVH), stroke, or hypoxic-ischemic injuries, were noted both preoperatively (n = 38, 44.7%) and postoperatively (n = 30, 35.3%). WMI was present in 26 of 85 preoperative scans (30.6%). New WMI was present on 21/85 postoperative scans (24.7%). Details on the type and severity of brain injury are included in Supplemental Table S1 (Online).

### Structural connectivity development

Network integration (global efficiency) and network segregation (transitivity and modularity) increased with postmenstrual age. This relationship was strongest for global efficiency, with 88% of subjects with both scans showing an increase between timepoints (n = 38/43). In univariate testing, gestational age at birth was associated with global efficiency (β_Interaction_ = 0.0048; SE = 0.0019; *p* = 0.014). Maternal education was not significantly associated with age-related change (β_Interaction_ = 0.00026; SE = 0.00015; *p* = 0.085) but was retained as a covariate. Sex was associated with age-related changes in modularity (β_Interaction_ = −0.029; SE = 0.009; *p* = 0.002), such that the age-related increase in modularity was more pronounced in male neonates than in females. Transitivity was not significantly associated with gestational age, sex, or maternal education level.

### Cardiac physiology is not a significant predictor of structural connectivity development

When assessing for differences in structural connectivity development by cardiac lesion (Figure 2), there were no differences found in the rate of growth of global efficiency (β_Interaction_ = −0.0009; SE = 0.0011; *p* = 0.418), transitivity (β_Interaction_ = −0.009; SE = 0.0021; *p* = 0.687), or modularity (β_Interaction_ = −0.003; SE = 0.011; *p* = 0.754) for neonates with SVP as compared to d-TGA.

**Figure 2.**
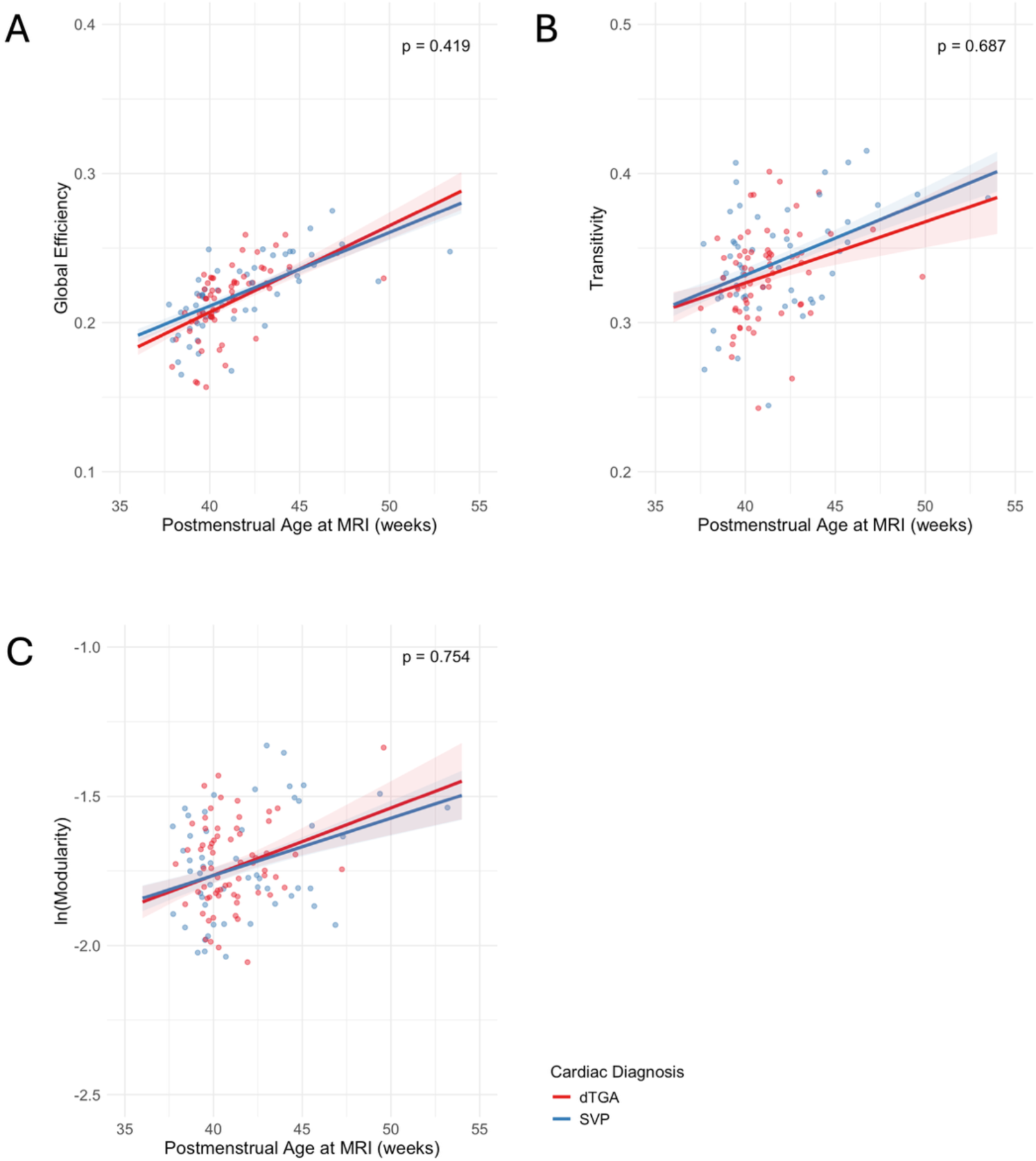
The effect of cardiac physiology on structural connectivity development. Linear mixed-effects linear models for graph metrics as a function of postmenstrual age at MRI scan, including global efficiency (A) with covariates gestational age and maternal education; transitivity (B); and modularity (C) with covariate sex. P-values shown are for the interaction between the cardiac diagnosis and time, which reflects the difference in groups in the rate of change of the tested graph metric.

### Brain injury significantly impairs network integration

When assessing for differences in structural connectivity development by the presence of any brain injury, including WMI, stroke, IVH, and hypoxic-ischemic injuries (Supplemental Figure S1 (online)), there were no differences found in the rate of growth of transitivity (β_Interaction_ = −0.0004; SE = 0.0019; *p* = 0.799) or modularity (β_Interaction_ = −0.026; SE = 0.010; *p* = 0.171). However, the rate of growth of global efficiency was significantly impaired in the presence of brain injury (β_Interaction_ = −0.0023; SE = 0.0001; *p* = 0.023). In linear mixed-effects models testing the effects of WMI (Figure 3) or stroke (Supplemental Figure S2 (online)) separately on network development, presence of WMI (β_Interaction_ = −0.003; SE = 0.001; *p* = 0.013), but not stroke (β_Interaction_ = 0.002; SE = 0.002; *p* = 0.168), was negatively associated with the rate of growth in Global Efficiency. There were no significant differences in network segregation by the presence of either WMI or stroke. Differences in structural connectivity development associated with intraventricular hemorrhage and hypoxic-ischemic injuries were not assessed, given their lower frequency during the perioperative period.

**Figure 3.**
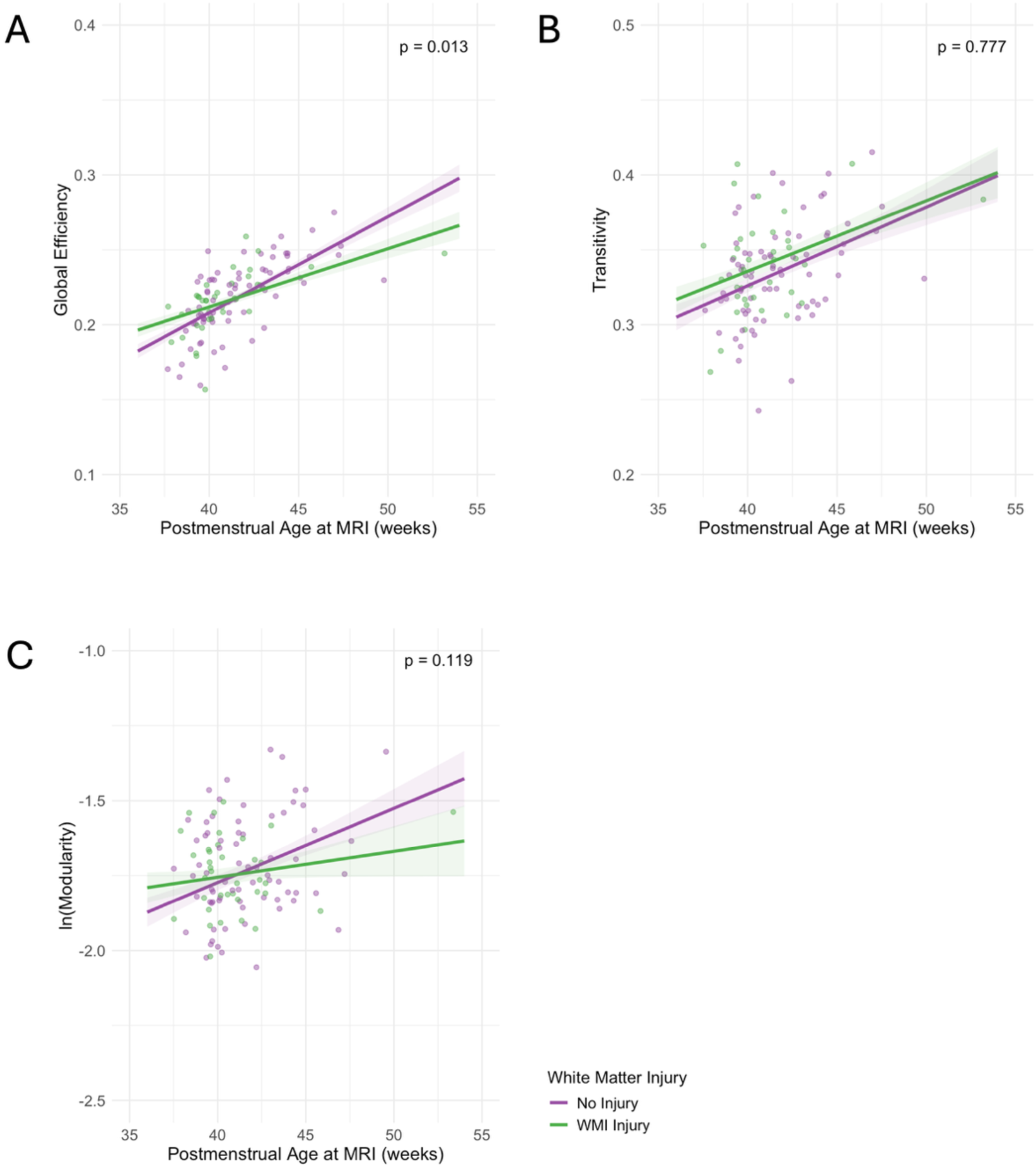
The effect of white matter injury on structural connectivity development. Linear mixed-effects linear models for global efficiency (A), including covariates gestational age and maternal education; transitivity (B); and modularity (C), including covariate sex. P-values shown are for the interaction between the presence of white matter injury and time, which reflects the difference in groups in the rate of change of the tested graph metric.

### Trajectory of network integration development during the perioperative period predicts early neurodevelopmental outcomes

Among infants eligible for follow-up, 82% (n= 50/61) completed at least one assessment with the BSID-III or −4 at 11.9 to 38.5 months corrected age during neurodevelopmental follow-up visits. The majority of subjects (n = 40, 80%) were tested more than once (Supplemental Figure S3 (online)). Results from univariate covariate testing are shown in Supplemental Table S2 (Online). After controlling for significant covariates, global efficiency trajectory (i.e., standardized slope) was significantly associated with cognitive (*p* = 0.001), language (*p* < 0.001), and motor (*p* = 0.008) scores (Table 2). For each 1-standard deviation increase in the predicted trajectory of global efficiency, there was an 8.2-point increase in BSID cognitive score (95% CI: 3.64-12.79), a 9.1-point increase in language scores (95% CI: 4.36-13.80), and a 5.7-point increase in motor scores (95% CI: 1.66-9.66).

**Table 2.**
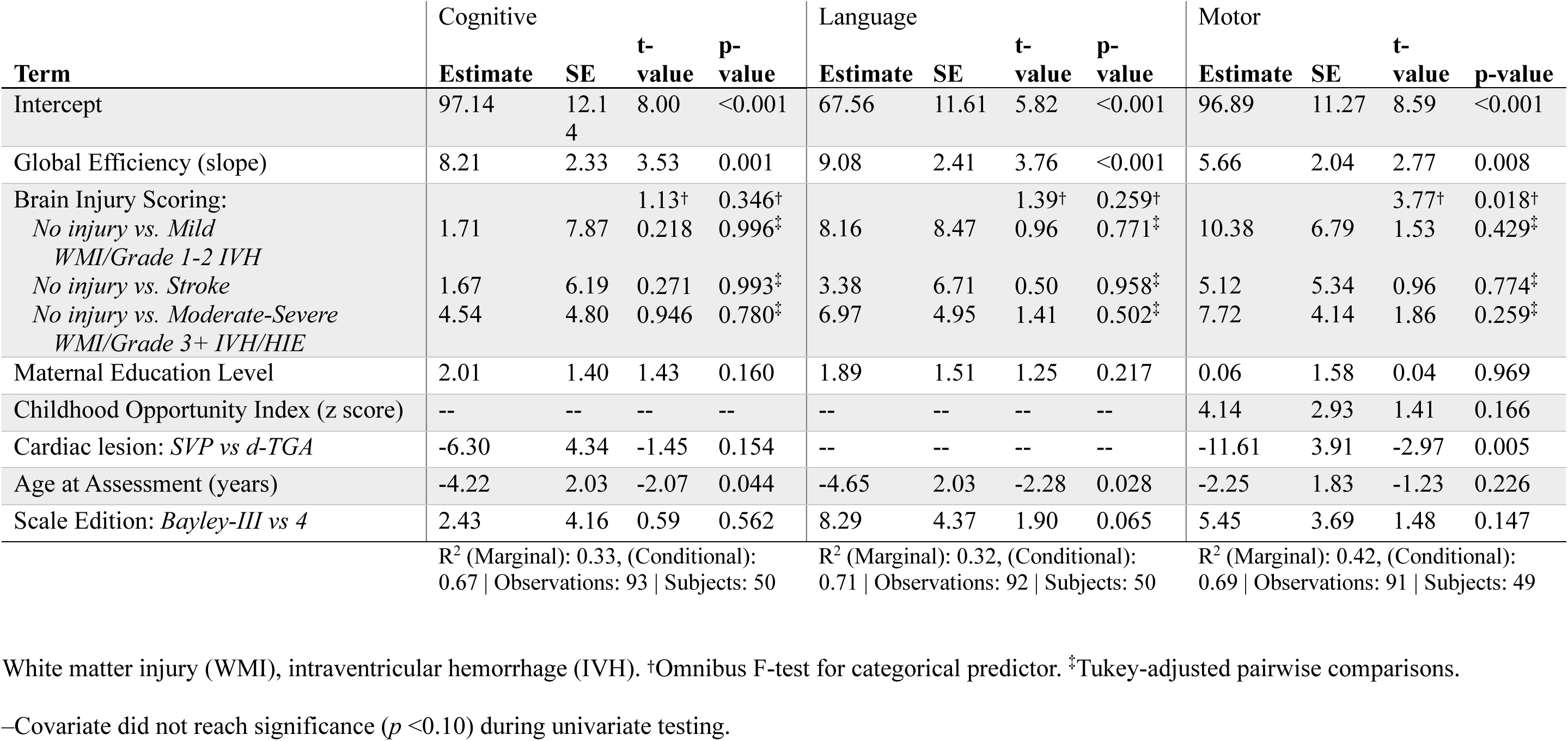
Multivariate linear mixed-effects model results for Bayley Scales of Infant & Toddler Development composite scores.

## Discussion

In this study, perioperative neural network integration, measured by global efficiency, increased with postmenstrual age. However, brain injury—particularly white matter injury (WMI)—attenuated this perioperative growth in global efficiency. Importantly, greater perioperative increases in global efficiency were associated with higher BSID scores across cognitive, motor, and language domains. Overall, these findings support the trajectory of neonatal network integration as a unique, early biomarker of neurodevelopmental outcomes in CHD.

### Cardiac physiology and network development

Contrary to our original hypothesis, cardiac physiology did not significantly affect the rate of neural network development. Neonates with single-ventricle physiology did not show slower perioperative increases in global efficiency or other network metrics than infants with d-TGA. The absence of a physiology-specific effect (d-TGA vs SVP) on perioperative network growth in our cohort contrasts with volumetric work suggesting lesion-specific growth differences (e.g., HLHS vs TGA)^17,18^. This difference highlights how brain size and brain network organization may capture distinct aspects of neurodevelopment. Our findings suggest that the topology of developing brain networks may also demonstrate a degree of resilience even in the setting of ongoing hypoxemia and altered brain growth.

### Brain injury and network integration

In contrast to cardiac physiology, perioperative brain injury significantly affected network development. Specifically, WMI was associated with slower growth of global efficiency during the perioperative period. WMI is the most common form of acquired brain injury in neonates with CHD and has been repeatedly associated with abnormalities in white matter microstructure and connectivity^2,19^. Our findings are consistent with prior work demonstrating disrupted connectome organization in CHD populations and the impact of WMI^10^. Our results extend these findings by demonstrating that WMI not only affects cross-sectional measures of connectivity but also impairs the trajectory of network development during the perioperative period. Importantly, rates of WMI appear to be declining in more contemporary cohorts of infants with CHD^20^. These findings highlight the importance of continued efforts to minimize perioperative brain injury, as even subtle disruptions to early network development may have downstream effects on neurodevelopment.

### Network development as a predictor of neurodevelopmental outcomes

Most importantly, we found that infants with larger perioperative increases in global efficiency had higher BSID cognitive, motor, and language scores across childhood. Identifying reliable early imaging predictors of neurodevelopmental outcomes in CHD has historically been challenging, and studies examining neonatal brain injury, particularly WMI, have reported heterogeneous results^17,20,21^. Ramirez et al. reported that neonatal global efficiency predicted motor outcomes at 30 months, but not earlier developmental scores, in a mixed cohort of infants with CHD and hypoxic-ischemic encephalopathy^11^. Our findings are unique in demonstrating that the trajectory of network integration during the neonatal perioperative period predicts cognitive, language, and motor outcomes across early childhood. To date, no other single neuroimaging biomarker has successfully predicted all neurodevelopmental outcome domains.

### The value of modeling developmental trajectories

One important difference between our study and much of the existing literature is the focus on modeling developmental trajectories rather than single timepoints. Neurodevelopmental challenges in children with CHD often emerge over time. Early motor delays may be detectable in infancy, followed by language difficulties in toddlerhood, while higher-order cognitive and executive deficits often become more apparent at school age as children as asked to do more^22^.

By modeling longitudinal changes in network efficiency rather than relying on a single neonatal timepoint, we were able to assess individual variation in early brain maturation and link these trajectories to developmental outcomes. This trajectory-based approach is likely a more sensitive marker of developmental risk than cross-sectional imaging findings alone. When we restricted the analysis to subjects with two complete imaging timepoints, our results were similar, which supports the robustness of our approach.

### Network resilience and plasticity

These findings raise the possibility that global efficiency may reflect a systems-level marker of neural resilience. Brain networks are highly dynamic and can reorganize after early injury or developmental disruption. Even in the setting of acquired brain injuries, alternative pathways may develop and support communication across brain regions. From this perspective, infants whose networks continue to develop efficiently during the perioperative period may retain the capacity to support strong cognitive, language, and motor outcomes. Network-level measures, such as global efficiency, may therefore capture aspects of brain development that are not reflected in focal injury measures or in volumetric brain measures alone.

### Clinical implications

Early identification of infants at highest risk for neurodevelopmental impairment remains a major challenge in CHD. Our findings suggest that perioperative gains in global efficiency may represent an early imaging marker associated with neurodevelopmental risk and could help identify infants who may benefit from closer developmental monitoring. The observed 8-point increase in BSID cognitive scores per standard deviation increase in the global efficiency trajectory is clinically meaningful, approaching the magnitude of differences associated with major perinatal risk factors such as prematurity^23^ or socioeconomic disadvantage^24^. Future multicenter studies are needed to determine the clinical utility of connectome measures as neuroimaging biomarkers. As neonatal MRI becomes increasingly incorporated into clinical care for infants with CHD, these measures may help inform individualized risk stratification and guide early developmental surveillance.

### Limitations

Our findings are strengthened by standardized perioperative imaging, longitudinal neurodevelopmental follow-up, and a two-stage analytic approach that derived subject-specific efficiency trajectories and linked them to BSID scores across multiple domains. However, there are some limitations. First, our data come from a single-center cohort, and not all infants underwent MRI at both perioperative time points. To incorporate infants with a single imaging dataset, we used a Bayesian mixed-effects framework to estimate subject-level trajectories of global efficiency. Although Bayesian mixed-effects modeling can estimate subject-specific slopes in this setting, these estimates may be less precise and may reflect partial pooling toward the population mean. Second, we assessed neurodevelopmental outcomes using BSID scores obtained between 1 and 4 years of age and from two test editions. This increased the available sample and allowed repeated measures to be incorporated, but assessments during early childhood may not fully capture later-emerging cognitive and executive difficulties that are common in children with CHD. Finally, because this was an observational cohort study, residual confounding from unmeasured differences in clinical severity, perioperative management, or socioeconomic context may have contributed to the associations we observed between network development and neurodevelopmental outcomes.

### Conclusion

In summary, perioperative structural connectome development is closely linked to early neurodevelopment in infants with critical CHD. While cardiac physiology did not significantly affect network development, perioperative brain injury—particularly white matter injury—was associated with slower growth in network integration. Most importantly, the trajectory of global efficiency development during the neonatal perioperative period predicted cognitive, language, and motor outcomes during early childhood. These findings suggest that early network development may serve as a sensitive biomarker of neurodevelopmental risk and resilience in this population.

## Author Contributions

LH, AK, DX, AS, EG, SP and PM provided substantial contributions to conception and design, acquisition of data, or analysis and interpretation of data. LH, SP and PM participated in drafting the article or revising it critically for important intellectual content. Final approval of the version to be published was provided by all authors.

## Consent Statement

This study was approved by the University of California, San Francisco Institutional Review Board. Written informed consent was obtained from parents or legal guardians of all participants.

## Data Sharing Statement

Deidentified participant data and analytic code are not publicly available at the time of preprint posting. Deidentified participant data and analytic code will be made available after peer-reviewed publication to researchers upon reasonable request, subject to a signed data-sharing agreement, as well as IRB approval or waiver as applicable, and approval by the corresponding author.

## Clinical Trial Registration

Not applicable; this observational cohort study did not prospectively assign participants to a health-related intervention.

---

**Figure S1.**
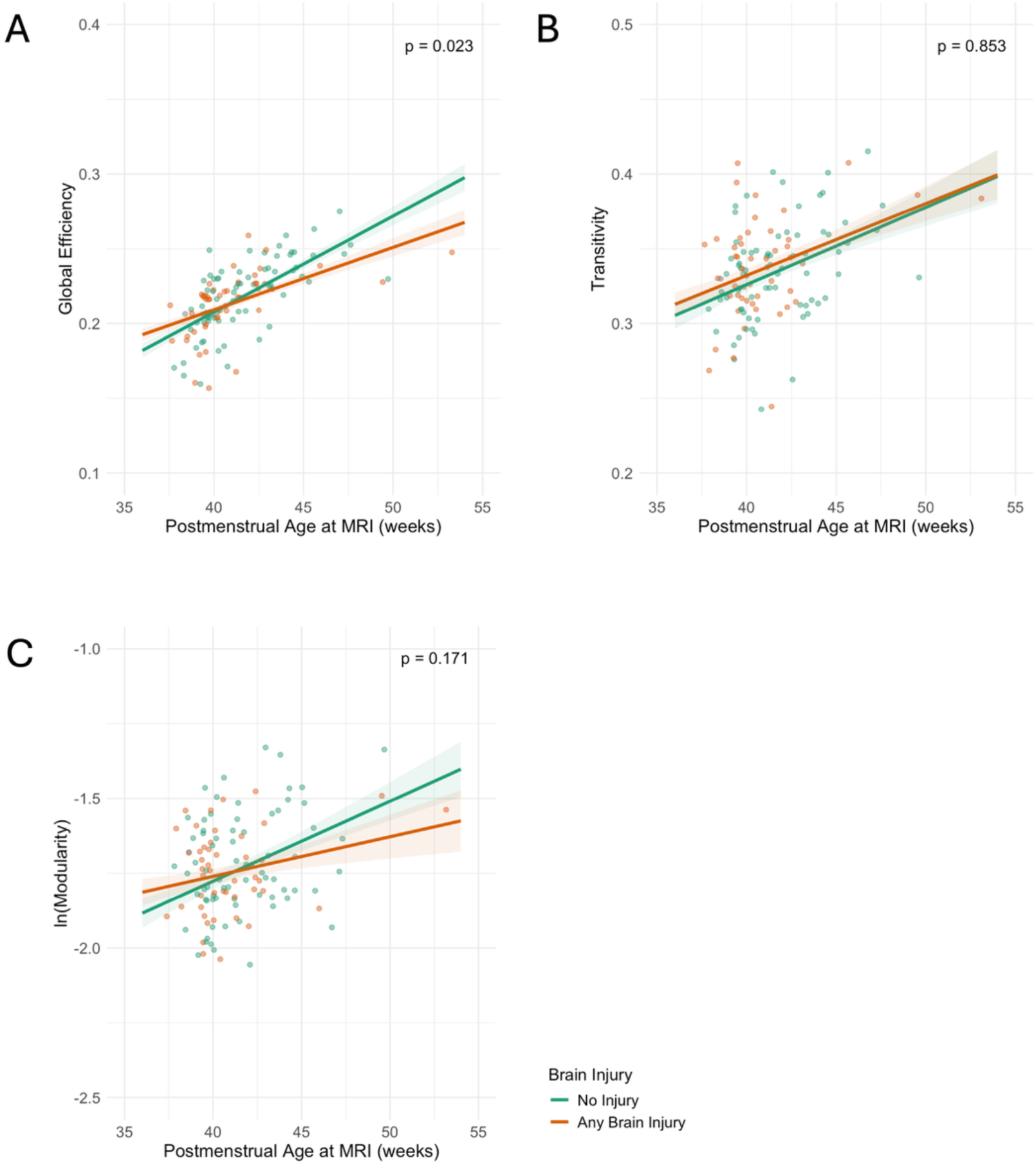
The effect of brain injury on structural connectivity development. Linear mixed-effects linear models for global efficiency (A), including covariates gestational age and maternal education; transitivity (B); and modularity (C), including covariate sex. Brain injury includes white matter injuries, intraventricular hemorrhage, stroke, and hypoxic-ischemic injuries. P-values shown are for the interaction between the presence of brain injury and time, which reflects the difference in groups in the rate of change of the tested graph metric.

**Figure S2.**
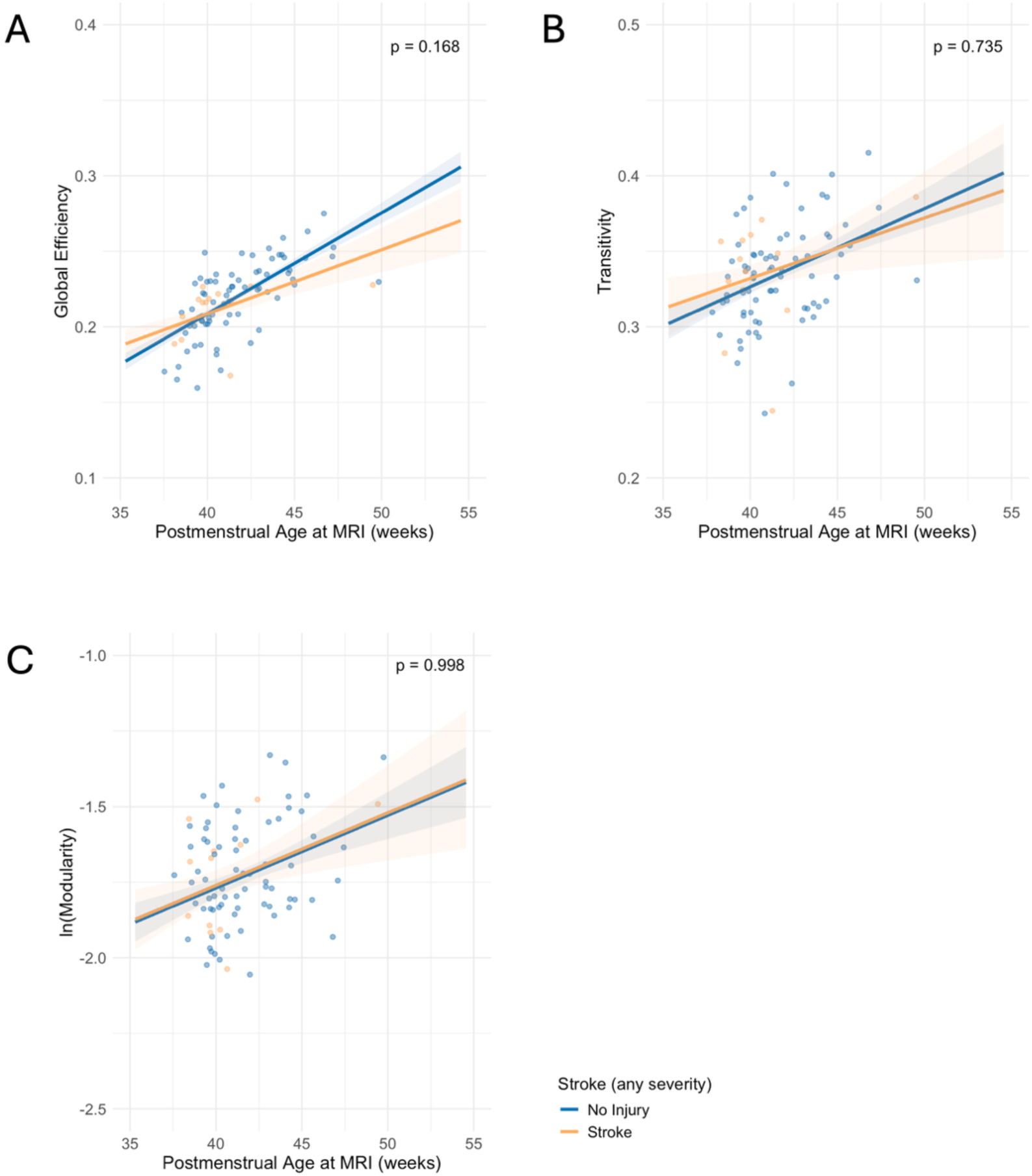
The effect of stroke on structural connectivity development. Linear mixed-effects linear models for global efficiency (A), including covariates gestational age and maternal education; transitivity (B); and modularity (C), including covariate sex. P-values shown are for the interaction between the presence of stroke and time, which reflects the difference in groups in the rate of change of the tested graph metric.

**Figure S3.**
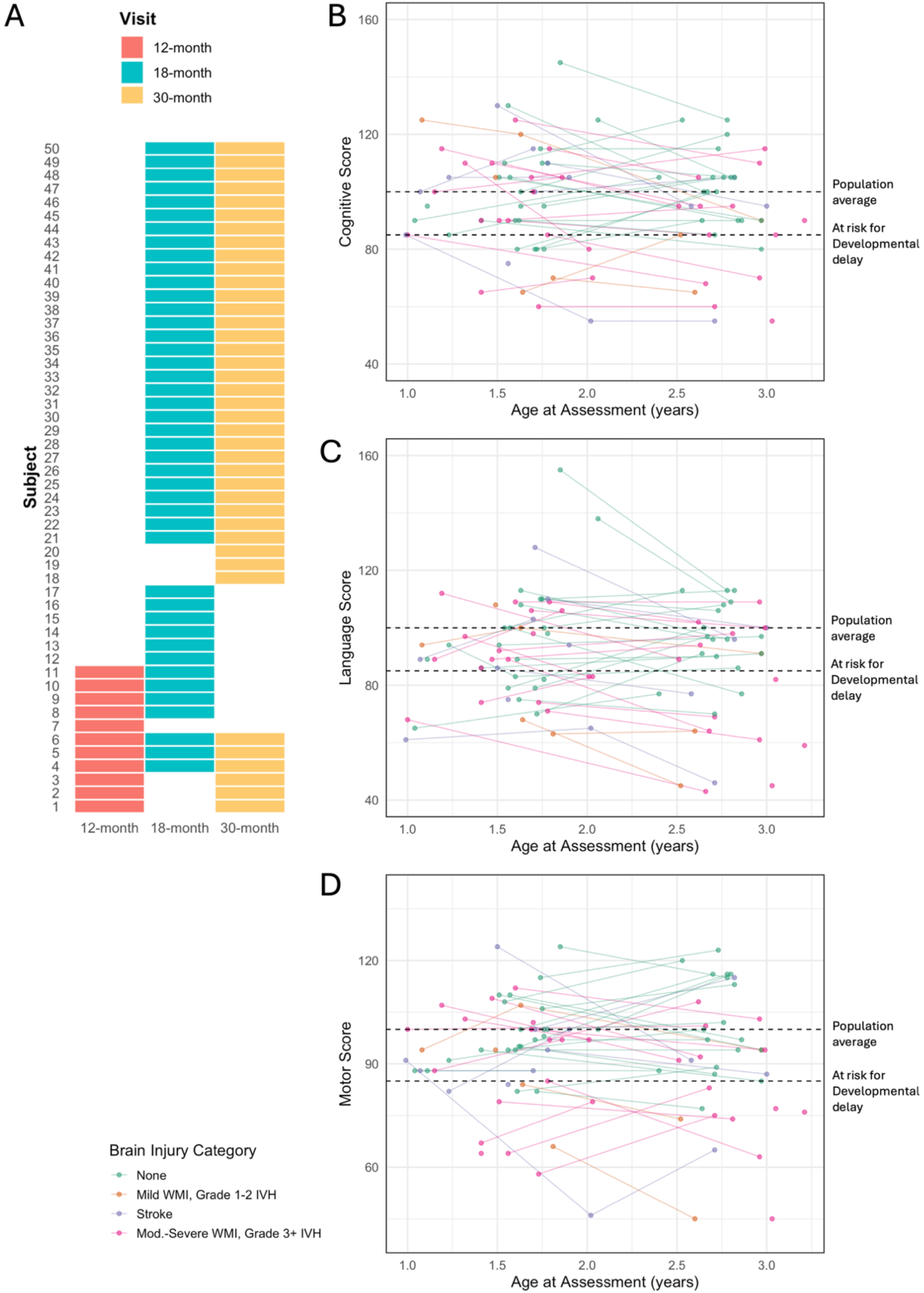
Neurodevelopmental follow-up testing. Distribution of follow-up visits completed by each subject (A). During the study period, the initial target visit shifted from 12 to 18 months to align with updated clinical practices. Score trajectories across follow-up visits for the Cognitive (B), Language (C), and Motor (D) subscales of the Bayley Scales of Infant & Toddler Development.

**Table S1.** Frequency of brain injury compared across the perioperative period.

|  | **Preoperative**<br>**Scan**<br>**(n=85)** | **Postoperative**<br>**Scan**<br>**(n=85)** | **p-**<br>**value<sup>1</sup>** |
| --- | --- | --- | --- |
| *Brain injury* |  |  | 0.21 |
| None | 47 (55.3) | 55 (64.7) |  |
| Any brain injury<sup>2</sup> | 38 (44.7) | 30 (35.3) |  |
| *White matter injury* |  |  | 0.49 |
| No injury | 59 (69.4) | 64 (75.3) |  |
| Mild injury (1-3 foci, all <2 mm) | 8 (9.4) | 8 (9.4) |  |
| Moderate injury (>3 foci or any foci >2 mm) | 17 (20.0) | 11 (12.9) |  |
| Severe injury (>5% of WM volume) | 1 (1.2) | 2 (2.4) |  |
| *Stroke* |  |  | 0.62 |
| No injury | 75 (88.2) | 77 (90.6) |  |
| Mild stroke (<1/3 vascular/parenchymal territory) | 8 (9.4) | 8 (9.4) |  |
| Moderate stroke (1/3 – 2/3) | 1 (1.2) |  |  |
| Severe stroke (>2/3) | 1 (1.2) |  |  |
| *Intraventricular hemorrhage* |  |  | <0.001 |
| No injury | 70 (82.4) | 83 (97.6) |  |
| Grade 1 | 3 (3.5) | 2 (2.4) |  |
| Grade 2 | 11 (12.9) | 0 (0.0) |  |
| Grade 3 | 1 (1.2) | 0 (0.0) |  |
| Grade 4 | 0 (0.0) | 0 (0.0) |  |
| *Hypoxic-ischemic injury* |  |  | 0.50 |
| None | 84 (98.2) | 85 (100) |  |
| Present | 1 (1.2) | 0 (0.0) |  |

<sup>1</sup>P-value from Fisher's exact test for the difference in prevalence (any injury vs. none) between the preoperative and postoperative scans for each injury type. <sup>2</sup>Includes white matter injury, intraventricular hemorrhage, stroke, and hypoxic-ischemic injury.

**Table S2.**
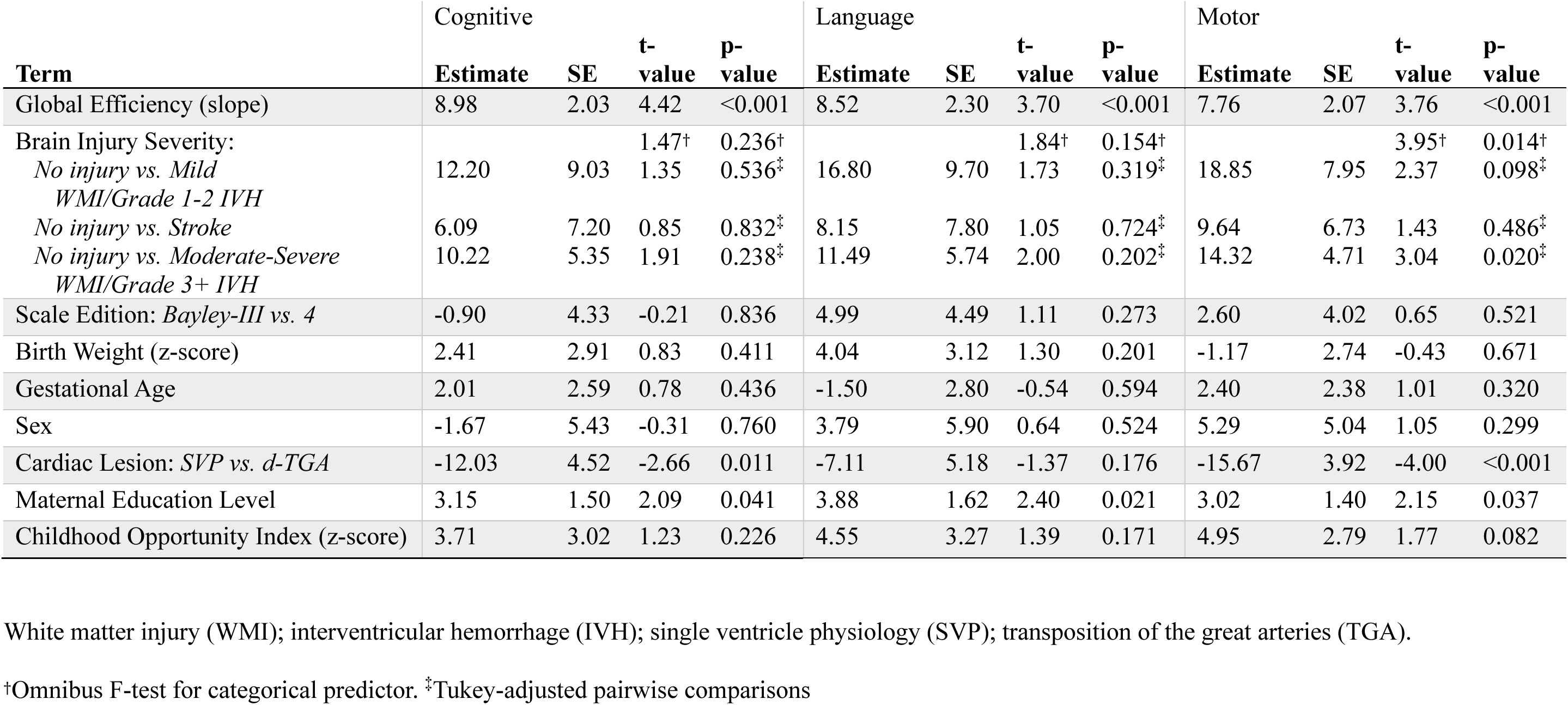
Univariate testing for Bayley Scales of Infant & Toddler Development composite scores.

## References

1 Sood, E. et al. Neurodevelopmental Outcomes for Individuals with Congenital Heart Disease: Updates in Neuroprotection, Risk-Stratification, Evaluation, and Management: A Scientific Statement from the American Heart Association. Circulation 149, e997–e1022 (2024).

2 Miller, S. P. et al. Abnormal Brain Development in Newborns with Congenital Heart Disease. N Engl J Med 357, 1928–1938 (2007).

3 Limperopoulos, C. et al. Brain Volume and Metabolism in Fetuses with Congenital Heart Disease: Evaluation with Quantitative Magnetic Resonance Imaging and Spectroscopy. Circulation 121, 26–33 (2010).

4 Phillips, K. et al. Neuroimaging and Neurodevelopmental Outcomes among Individuals with Complex Congenital Heart Disease. JACC 82, 2225–2245 (2023).

5 Dijkhuizen, E. I. et al. Early Brain Magnetic Resonance Imaging Findings and Neurodevelopmental Outcome in Children with Congenital Heart Disease: A Systematic Review. Developmental Medicine & Child Neurology 65, 1557–1572 (2023).

6 Ortinau, C. M. et al. Optimizing Neurodevelopmental Outcomes in Neonates with Congenital Heart Disease. Pediatrics 150 (2022).

7 Peyvandi, S., Latal, B., Miller, S. P. & McQuillen, P. S. The Neonatal Brain in Critical Congenital Heart Disease: Insights and Future Directions. NeuroImage 185, 776–782 (2019).

8 Hagmann, P., Grant, P. E. & Fair, D. A. Mr Connectomics: A Conceptual Framework for Studying the Developing Brain. Frontiers in Systems Neuroscience Volume 6 - 2012 (2012).

9 Ball, G. et al. Rich-Club Organization of the Newborn Human Brain. Proc Natl Acad Sci U S A 111, 7456–7461 (2014).

10 Feldmann, M. et al. Delayed Maturation of the Structural Brain Connectome in Neonates with Congenital Heart Disease. Brain Commun 2, fcaa209 (2020).

11 Ramirez, A. et al. Neonatal Brain Injury Influences Structural Connectivity and Childhood Functional Outcomes. PLoS One 17, e0262310 (2022).

12 Bayley, N. Bayley Scales of Infant and Toddler Development 3 edn(Harcourt Assessment, 2006).

13 Bayley, N. & Aylward, G. P. Bayley Scales of Infant and Toddler Development 4 edn(Pearson, 2019).

14 Tymofiyeva, O. et al. Towards the “Baby Connectome”: Mapping the Structural Connectivity of the Newborn Brain. PLoS One 7, e31029 (2012).

15 Dimitropoulos, A. et al. Brain Injury and Development in Newborns with Critical Congenital Heart Disease. Neurology 81, 241–248 (2013).

16 Acton, B. V. et al. Overestimating Neurodevelopment Using the Bayley-Iii after Early Complex Cardiac Surgery. Pediatrics 128, e794–800 (2011).

17 Andropoulos, D. B. et al. Brain Immaturity Is Associated with Brain Injury before and after Neonatal Cardiac Surgery with High-Flow Bypass and Cerebral Oxygenation Monitoring. J Thorac Cardiovasc Surg 139, 543–556 (2010).

18 Peyvandi, S. et al. The Association between Cardiac Physiology, Acquired Brain Injury, and Postnatal Brain Growth in Critical Congenital Heart Disease. J Thorac Cardiovasc Surg 155, 291–300.e293 (2018).

19 Licht, D. J. et al. Brain Maturation Is Delayed in Infants with Complex Congenital Heart Defects. J Thorac Cardiovasc Surg 137, 529–536; discussion 536-527 (2009).

20 Peyvandi, S. et al. Neonatal Brain Injury and Timing of Neurodevelopmental Assessment in Patients with Congenital Heart Disease. J Am Coll Cardiol 71, 1986–1996 (2018).

21 Beca, J. et al. New White Matter Brain Injury after Infant Heart Surgery Is Associated with Diagnostic Group and the Use of Circulatory Arrest. Circulation 127, 971–979 (2013).

22 Sanz, J. H. et al. Trajectories of Neurodevelopment and Opportunities for Intervention across the Lifespan in Congenital Heart Disease. Child Neuropsychol 29, 1128–1154 (2023).

23 Anderson, P. J. & Doyle, L. W. Cognitive and Educational Deficits in Children Born Extremely Preterm. Semin Perinatol 32, 51–58 (2008).

24 Hackman, D. A., Farah, M. J. & Meaney, M. J. Socioeconomic Status and the Brain: Mechanistic Insights from Human and Animal Research. Nat Rev Neurosci 11, 651–659 (2010).

